# Unique predictors of intended uptake of a COVID-19 vaccine

**DOI:** 10.1101/2020.12.11.20235838

**Authors:** Robert P. Lennon, Meg L. Small, Rachel A. Smith, Lauren J. Van Scoy, Jessica G. Myrick, Molly A. Martin, Data4Action Research Group

## Abstract

**Introduction:** An effective vaccine for COVID-19 is only of value if the public has confidence in taking it. There is little data on COVID-19-specific vaccine confidence or its determinants in the United States. The objective of this study was to determine public confidence in a COVID-19 vaccine.

**Methods:** A cross-sectional survey of Pennsylvanian adults, August-October, 2020, to identify their likelihood of taking an approved, no-cost coronavirus vaccine, general vaccine acceptance, and sociodemographic traits to identify predictors of vaccine acceptance.

**Results:** Of the 950 respondents, 55% were “very likely”, 20% “somewhat likely”, 14% “unsure”, 4% “somewhat unlikely”, and 7% “very unlikely” to take a coronavirus vaccine, even though 70% had taken the flu vaccine since September 2019. The strongest predictors of vaccine acceptance were trust in the system evaluating vaccines and perceptions of local COVID-19 vaccination norms. The strongest predictors of negative vaccine intentions were worries about unknown side-effects and positive attitudes toward natural infection. Sociodemographic factors, political views, and religiosity did not predict vaccine intentions.

**Conclusions:** Fewer adults intend to take a coronavirus vaccine than currently take the flu vaccine. To overcome coronavirus vaccine hesitancy, information campaigns to reinforce positive predictors and overcome negative predictors are indicated.

## INTRODUCTION

An effective SARS-CoV-2 vaccine to stop the spread of COVID-19 is only of value if the public has confidence in taking it.^1^ Concerns about public refusal to accept a COVID-19 vaccine are widespread^2^ and have raised calls for drastic measures to ensure public confidence in COVID-19 vaccines developed.^3^ Pre-COVID-19, the most important determinants of vaccine uptake globally were confidence in vaccines and trust in health-care workers, with lesser variations in confidence by science education, age, and gender, and little variation by income or religion.^4^

There is little data on COVID-19-specific vaccine confidence or its determinants in the United States (U.S.). A survey of 672 U.S. adults in May, 2020, found 67% would accept a COVID-19 vaccine, with variations in confidence by demographic status.^5^ A July, 2020 survey of 1,971 U.S. adults found associations between probability of choosing a COVID-19 vaccine and perceived vaccine efficacy, side effect profile, which institution endorsed it, and the national origin of the vaccine.^6^ A more recent public poll in September, 2020, found only 51% of U.S. adults would definitely or probably get a COVID-19 vaccine – down from 72% from the same poll in May, 2020.^7^ That poll identified safety concerns as a primary reason for hesitancy, including concerns about both the development process and approval process of a COVID-19 vaccine, and also found differences in acceptance by race, ethnicity, and political affiliation.^7^

This drop in intended vaccine uptake is concerning and may represent a shift in positive and negative predictors of vaccine confidence. The objective of this study was to explore COVID-19 vaccine confidence in light of these apparent changes.

## METHODS

### Population

This manuscript reports data from a longitudinal cohort survey at time 1, administered either online or by telephone, of adults in Centre County, Pennsylvania (PA), collected between August and October, 2020. Centre County, PA has approximately 150,000 permanent residents, with an academic year increase in population of approximately 40,000 undergraduate students enrolled to the Pennsylvania State University, University Park campus. Participants were recruited between May and September, 2020 through employers, local government, news coverage and word-of-mouth, with eligibility requirements of age 18 years or older and residence in Centre County, PA. Of 1,590 overall enrollees, 986 were invited to complete this survey, and 950 completed it (96% completion rate; no missing data). This study was approved by the Pennsylvania State University IRB.

### Measures

Our primary outcome was COVID-19 vaccination intention. Participants answered, “*If an FDA-approved vaccine to prevent COVID-19 were available today at no cost to you, how likely would you be to get vaccinated?”* on a 5-point scale (1=*very unlikely*, 5=*very likely*).

Independent variables included vaccination attitudes, norms, efficacy, and past behavior.^8^ Positive attitudes toward vaccination (3 items; α=.77) and immunity through natural infection (versus vaccination) (3 items; α=.77) were measured on a 5-point scale (1=*strong disagree*, 5=*strongly agree*). Perceived vaccine uptake norms were measured by asking participants to estimate what percentage of people living in the county would get the coronavirus vaccine (1m item, 0-20%; 21-40%; 41-60%; 61-80%, 81-100%). Efficacy was measured by asking participants if they worry about unknown side-effects of vaccines (1 item, 1=*strong disagree*, 5=*strongly agree*). Past behavior was measured by asking participants whether they received a flu vaccine since September 1, 2019 (yes, no, wanted to get the flu vaccine, but were unable to for medical reasons; for regression analysis, the latter categories were combined).

Trust has also been associated with vaccine intentions and behaviors, and was measured as whether the respondent trusts the current system for evaluating the safety of coronavirus vaccines (1 item, 1=*strong disagree*, 5=*strongly agree*), and general vaccine cynicism (2 items, *r* = .63, *p*<.001; e.g., *Authorities promote vaccination for financial gain, not for people’s health*). Sociodemographic variables of education, financial standing, political viewpoint (1=*very conservative*, 7=*very liberal*), and religiosity were also measured.

### Statistical Analysis

Descriptive statistics were used to describe quantitative data. Multivariate ordinal regression was used to model predictors of vaccine intention. Statistical analysis was completed using SPSS statistical software version 25.

## RESULTS

The 950 respondents were characteristically middle age (49 years, *SD*= 16.32), highly educated (87% Bachelor’s degree or higher), employed (74%), non-Hispanic (95%), Caucasian (94%), women (68%).

Just over half (55%) reported that they are “very likely,” to take a coronavirus vaccine, while 20% are “somewhat likely,” 14% are “unsure,” 4% are “somewhat unlikely,” and 7% are “very unlikely” to take a coronavirus vaccine, even though 70% had taken the flu vaccine this season.

Multivariate ordinal regression was used to evaluate predictors of vaccine intention (Table 1). The strongest predictors of positive vaccine intentions were trust in the vaccine and perceived county COVID-19 vaccination norms; positive attitudes toward vaccines generally and having received the flu vaccine last year were lesser positive predictors. The strongest predictor of negative vaccine intention, by far, was worries about unknown side-effects; positive attitudes toward natural infection (over vaccination) was a lesser negative predictor. Sociodemographic status, political views, and religiosity did not predict vaccine intentions.

**Table 1.** Multivariate ordinal regression model of predictors of coronavirus vaccination intention with qualitative thematic commentary.

| Variable | Estimate (SE) | Wald | Free-text illustrations of COVID-19-specific hesitancy |
| --- | --- | --- | --- |
| Trust in system evaluating coronavirus vaccine | .57 (.06) | **78.66\*\*\*** | *“I fear the long-term safety and efficacy of a rushed vaccine and think that safeguards may be ignored. . .”*<br>*“I think the FDA is being forced to rush things.”*<br>*“I am very pro-vaccine, but I am also extremely concerned that political pressures will affect the appropriate vaccine timelines.”*<br>*“It seems there is a lack of transparency in how many of the trials are being carried out and evaluated for safety. . .”* |
| Vaccine side-effect | -.41 (.06) | **46.86\*\*\*** |  |
| Perceived Norm | .54 (.08) | **46.3\*\*\*** |  |
| Positive attitudes toward Vaccine | .44 (.14) | **10.34\*\*** |  |
| Received flu vaccine in past year<sup>†</sup> | .21 (.08) | **7.69\*\*** |  |
| Positive attitudes toward Natural Infection | -.22 (.10) | **4.69\*** |  |
| Vaccine cynicism | -.22 (.13) | 2.72 |  |
| Religiosity | -.08 (.07) | 1.46 |  |
| Financial standing | .02 (.05) | 0.17 |  |
| Education | .09 (.07) | 1.53 |  |
| Liberal politics | .01 (.06) | 0.04 |  |
| Pseudo $R^2$ | .39 | | |

Boldface indicates statistical significance (\* $p < 0.05$ , \*\* $p < 0.01$ , \*\*\* $p < 0.001$ )
<sup>†</sup> For this regression, answers of, “no” and “could not for medical reasons” were combined.

Free-text participant responses were used to illustrate quantitative results.

## DISCUSSION

Our data identify a difference between confidence in vaccines in general and confidence in a COVID-19 vaccine, which stems from a primary distrust in the system evaluating the coronavirus vaccine. Free-text responses suggest that those unlikely to take a COVID-19 vaccine are worried that the speed of development indicates a lack of integrity in developing the vaccine, and that politicization of oversight agencies reduces their trustworthiness. This differs from general pre-COVID vaccine hesitancy, which primarily stemmed from negative vaccine beliefs, largely based on misconceptions.^9^ Whereas, historically, the primary emphasis for overcoming vaccine hesitancy was to overcome misconceptions, for COVID-19, these data suggest the primary emphasis must be on reassuring the public that the integrity of the system for evaluating the vaccine was maintained. This confirms public polling showing distrust in both the development and approval process for the coronavirus vaccine,^7^ and supports prior editorial recommendations for instituting a policy of radical transparency in the COVID-19 vaccine development process.^3^

### Limitations

Limitations to our study include that it is a single point in a cohort study; results at this time may not be generalizable over time. It is also limited by its narrow population sample, which was primarily well educated, Caucasian females.

Strengths of our study include a large sample size compared to prior US COVID-19 studies and timeliness. Capturing intent closer to the potential time of COVID-19 vaccine release is more likely to reflect actual uptake than older studies.

While further research is needed to confirm the generalizability of our results, given their concordance with the national Pew sample,^7^ it is highly likely that fewer US adults intend to take a coronavirus vaccine than currently take the flu vaccine. Further, given changes in positive and negative predictors, to overcome coronavirus vaccine hesitancy, practices, campaigns and policies may need to focus on reinforcing vaccine safety and development integrity over more general historic concerns.

## Conclusions

These results are directly relevant to the COVID-19 pandemic. Changing COVID-19 vaccine uptake intent and predictors of intent warrants changes to how COVID-19 public health vaccination campaigns are designed. For messages and messengers to effectively reinforce public uptake of a COVID-19 vaccine, they must emphasize the safety of the vaccine and the integrity of the vaccine development process as priorities.

## Data Availability

Data is not available at this time.

## Contributors

The authors thank the members of the D4A Action Research Group: Dee Bagshaw, Clinical & Translational Science Institute, Nita Bharti, Dept. of Biology and the Huck Institutes of the Life Sciences, Cyndi Flanagan, Clinical Research Center, Matthew Ferrari, Dept. of Biology & Huck Institutes of the Life Sciences, Thomas Gates, Social Science Research Institute, Margeaux Gray, Dept. of Biobehavioral Health, Suresh Kuchipudi, Animal Diagnostic Lab, Vivek Kapur, Dept. of Animal Science and the Huck Institutes of Life Sciences, Stephanie Lanza, Dept. of Biobehavioral Health and the Prevention Research Center, James Marden, Dept. of Biology & Huck Institutes of the Life Sciences, Susan McHale, Dept. of Human Development and Family Studies and the Social Science Research Institute, Glenda Palmer, Social Science Research Institute, Andrew Read, Depts. of Biology and Entomology, and the Huck Institutes of the Life Sciences, Connie Rogers, Dept. of Nutritional Sciences and the Huck Institutes of the Life Sciences, Meg Small, Prevention Research Center, Rachel Smith, Dept. of Communication Arts and Sciences and the Huck Institutes of the Life Sciences, Charima Young, The Penn State Office of Government and Community Relations.

## CRediT author statement

**Robert Lennon:** Methodology, Formal analysis, Writing – Original Draft, Writing – Review and Editing **Meg Small:** Conceptualization, Methodology, Formal analysis, Investigation, Writing – Review and Editing **Rachel Smith:** Conceptualization, Methodology, Validation, Formal analysis, Investigation, Writing – Original Draft, Writing – Review and Editing, Supervision, Funding Acquisition **Lauren Van Scoy:** Methodology, Writing – Review and Editing **Jessica Myrick:** Conceptualization, Methodology, Formal analysis, Investigation, Writing – Review and Editing **Molly Martin:** Conceptualization, Methodology, Formal analysis, Investigation, Writing – Review and Editing **Data4Action Research Group:** Conceptualization, Methodology, Resources, Project Administration, Funding Acquisition, Writing – Review and Editing

